# Low-intensity repetitive transcranial magnetic stimulation is safe and well tolerated by people living with MS - outcomes of the phase I randomised controlled trial (TAURUS)

**DOI:** 10.1101/2024.02.29.24303579

**Authors:** Phuong Tram Nguyen, Amin Zarghami, Kalina Makowiecki, Natasha Stevens, Chigozie Ezegbe, Kain Kyle, Chenyu Wang, Linda Ly, Katie De La Rue, Mark R Hinder, Lewis Johnson, Jennifer Rodger, Samantha Cooper, Carlie L Cullen, Michael Barnett, Kaylene M Young, Bruce V Taylor

**Author notes:** These authors contributed equally to this study (Phuong Tram Nguyen, Amin Zarghami, Kalina Makowiecki). Senior and corresponding authors, Bruce V Taylor Menzies Institute for Medical Research, University of Tasmania, 17 Liverpool St, Hobart, TAS 7000, Australia., Kaylene M Young Menzies Institute for Medical Research, University of Tasmania, 17 Liverpool St, Hobart, TAS 7000, Australia.

## Abstract

**Background:** Low-intensity repetitive transcranial magnetic stimulation (rTMS), delivered as a daily intermittent theta burst stimulation (iTBS) for 4 consecutive weeks, increases the number of new oligodendrocytes in the adult mouse brain. rTMS holds potential as a remyelinating intervention for people with MS.

**Objective:** Primarily to determine the safety and tolerability of our rTMS protocol in people with MS. Secondary objectives included feasibility, blinding, and an exploration of changes in magnetic resonance imaging (MRI) metrics, patient reported outcome measures (PROMs) and cognitive or motor performance.

**Methods:** A randomised (2:1), placebo controlled, single blind, parallel group, phase 1 trial of 20 rTMS sessions (600 iTBS pulses per hemisphere; 25% maximum stimulator output), delivered over 4-5 weeks, with the coil positioned at 90° (n=7, ‘sham’) or 0° (n=13, active rTMS).

**Results:** Five adverse events (AEs) including one serious AE reported. None were related to treatment. Protocol compliance was high (85%) and blinding successful. Within participant MRI metrics, PROMs and cognitive or motor performance were unchanged over time.

**Conclusion:** 20 sessions of rTMS is safe and well tolerated in a small group of people with MS. The study protocol and procedures are feasible. Improvement of sham is warranted before further investigating safety and efficacy.

**Trial registration:** Australian New Zealand Clinical Trials Registry (ACTRN12619001196134). https://www.anzctr.org.au/Trial/Registration/TrialReview.aspx?id=378010&isReview=true

## Introduction

Remyelination therapies remain a critical unmet need and research priority for people with multiple sclerosis (MS).^1^ In healthy and demyelinated mice, optogenetic and chemogenic approaches increase neuronal activity and myelin addition,^2^ however, these technologies cannot be used clinically. Learning can enhance remyelination in activated CNS region/s^3^ but is impractical for MS, when lesions are multifocal. By contrast, repetitive transcranial magnetic stimulation (rTMS) can non-invasively modulate neuronal activity.^4^ 28 consecutive daily sessions of low-intensity rTMS (LI-rTMS), delivered as an intermittent theta burst stimulation (iTBS), can increase the survival and maturation of new oligodendrocytes in the cortex of adult mice.^5^ It is unknown whether rTMS can also promote remyelination in humans.

rTMS is generally well-tolerated^6^ and can be well tolerated by people with MS.^7–16^ Our rTMS intervention differs from previous studies in several ways: we use a circular electromagnetic coil to deliver a diffuse stimulation and target a broad cortical area; rTMS is delivered at 20 intervention visits, and at a lower stimulation intensity. We conducted the present clinical trial to primarily evaluate safety and tolerability, and the promise of rTMS for promoting remyelination in people with MS. We also evaluated the feasibility of our randomisation, blinding, magnetic resonance imaging (MRI) and electronic patient reported outcome measure procedures for use in future phase II trials.

## Materials and Methods

### Study design

The low-intensity repetitive transcranial magneTic brAin stimUlation foR people living with mUltiple Sclerosis (TAURUS) trial is a phase I, randomised, placebo (‘sham’) controlled, single blind, parallel group trial, carried out according to the published trial protocol.^17^

This study was approved by the Human Research Ethics Committee, University of Tasmania (17899), and carried out in accordance with the World Medical Association’s Declaration of Helsinki, International Conference of Harmonisation Good Clinical Practice (ICHGCP) and The Australian Code for the Responsible Conduct of Research, 2018 (The Code). This trial was registered on the Australian New Zealand Clinical Trial Registry (ANZCTR) (ACTRN12619001196134).

A consumer and community reference committee (C&CRC) (n=14) provided advice on study design and feasibility, plain language and accessibility, participant information sheets, recruitment, promotional materials, levels of reimbursement and communication of trial results to the MS community. A concern of the C&CRC was the feasibility of people with MS completing 20 intervention visits.

### Participants

Participant eligibility criteria are detailed in our protocol.^17^ Participants were recruited at the Menzies Institute for Medical Research, Clinical Research Facility from 14 January 2021 to 27 July 2021. Thirty-six participants were screened. Twenty participants were eligible and gave written, informed consent to participate (**Figure 1**). **Table 1** provides baseline demographic data and the clinical characteristics of included participants.

**Figure 1.**
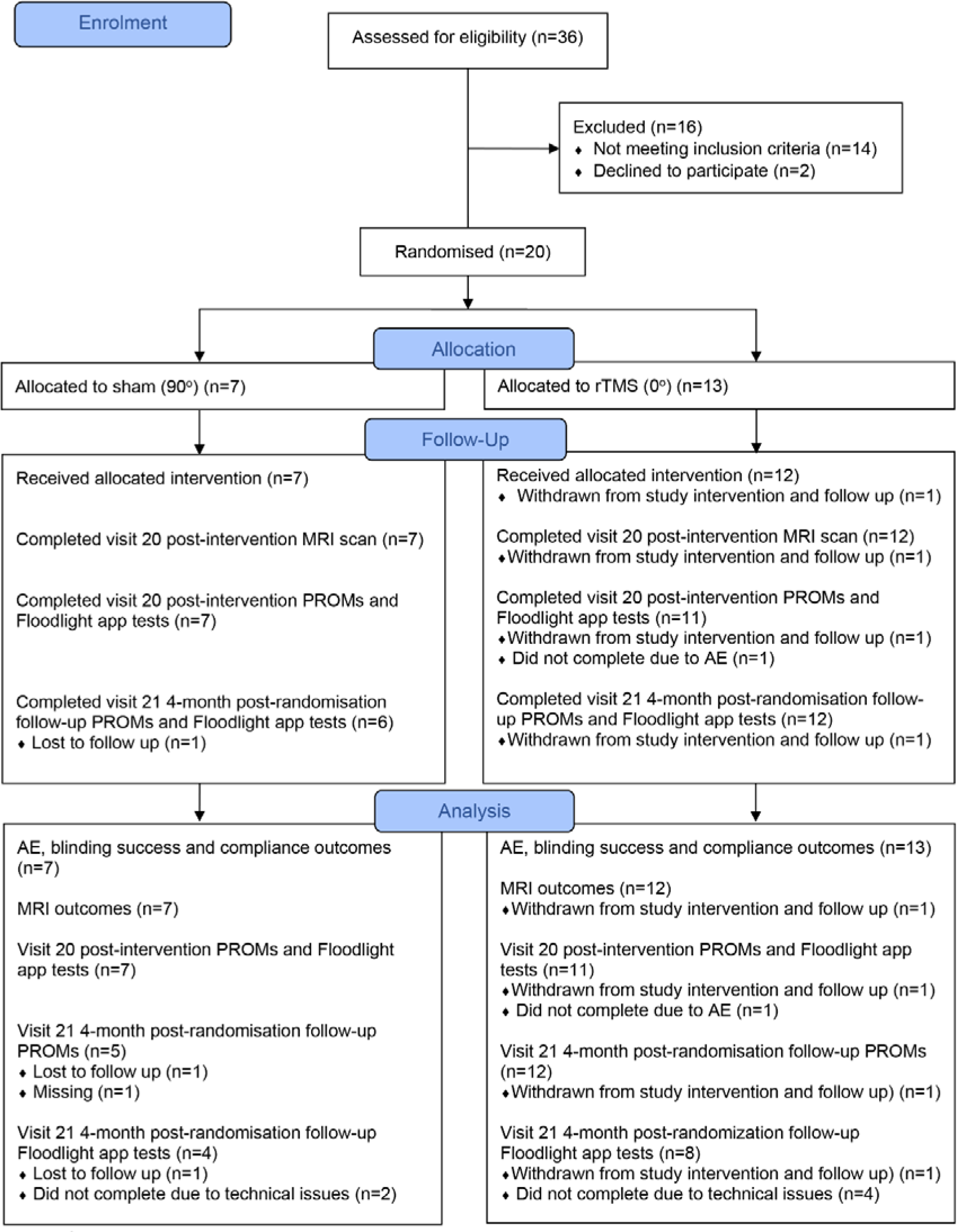
CONSORT flowchart showing the number of participants that were: screened, recruited, randomized to intervention, completed the intervention and follow-up, and were analysed.

**Table 1.** Baseline demographic and clinical characteristics.

|  | rTMS coil orientation |  |
| --- | --- | --- |
|  | Sham (90°)<br>n = 7 | rTMS (0°)<br>n = 13 |
| Sex assigned at birth, n (%) |  |  |
| Male | 2 (28.6%) | 4 (30.8%) |
| Female | 5 (71.4%) | 9 (69.2%) |
| Age [years], mean (SD) | 52 (± 4.3) | 52.8 (± 12.3) |
| MS duration [years], mean (SD) | 12.1 (± 9.4) | 15.2 (± 10.0) |
| EDSS score at baseline, median (IQR) | 2.5 (1.5 – 5) | 2.5 (2 – 3.5) |
| MS type, n (%) |  |  |
| Relapsing-remitting MS | 6 (85.7%) | 8 (61.5%) |
| Primary progressive MS | 0 (0.0) | 2 (15.4%) |
| Secondary progressive MS | 1 (14.3%) | 3 (23.1%) |

EDSS = Expanded Disability Status Scale; IQR = interquartile range; SD = standard deviation.

Participants were not required to stop concomitant MS treatments. Participants were followed up until the last participant’s final visit on 21 December 2021.

### Randomisation and blinding

Randomisation was performed immediately prior to intervention visit 1 by an unblinded researcher using an automated, real-time, central web-based REDCap system.^17^ For allocation concealment, we used a block size of 6, with a ratio of 1:2 allocation for the sham (90°) (n=7) and rTMS (0°) groups (n=13), stratified by sex.

Participants were blind to treatment allocation and researcher blinding was as described.^17^

### Interventions

rTMS was delivered using a Magstim Rapid2 (Magstim Rapid2 part numbers: 3012-00 Rapid2 Mainframe – 50Hz; 3013-00 Single Power Supply Unit; 3022-00 User Interface; Magstim Ltd, Whitland, UK) and Magstim 90mm standard circle coil (model: 3193-00, Magstim Ltd, Whitland, UK). The stimulation intensity was set at 25% maximum stimulator output and parameters set for iTBS pattern as described.^17^ rTMS was delivered at each of the 20 onsite intervention visits (≤ 5d per week for 4-5 consecutive weeks), allowing for a maximum gap of 3 consecutive days between intervention visits for intervention compliance.

For the rTMS (0°) group, rTMS was delivered according to our protocol.^17^ The plane of the circular coil was positioned tangential to the scalp, the central hole was over the target mark (2 cm lateral from the vertex), and the coil handle pointed backwards at 45° to the scalp midline. Stimulation was applied to a broad cortical area, including frontal and parietal regions, consistent with our preclinical protocol.^5^ For the sham (90°) group, the coil was tilted 90° relative to the scalp, such that the edge profile touched the scalp at the target location. Commercial circular sham coils are not available for this rTMS device. Prior to the development of sham figure-of-eight rTMS coils, it was common to position the rTMS coil at 90° for the sham group, to preserve the “clicking sound” associated with the intervention and significantly reduce (but not abolish) the stimulation.^18, 19^ As sham (90°) is not a no stimulation control, we evaluated within-participant changes from baseline in sham (90°) and rTMS (0°) groups, or compared with *a priori* criteria, to assess safety and tolerability outcomes.

### Outcomes

The primary outcome was incidence of treatment-emergent adverse events (AEs) and serious AEs (SAEs) experienced between intervention visit 1 and 20. A difference of 10% between groups would be considered clinically significant.

Secondary outcomes included: evaluation of AEs and SAEs at the 4-month post-randomisation follow up; protocol compliance; blinding success; changes in MRI, PROMs and cognitive or motor assessments.

#### Compliance

was assessed as the proportion of participants in the sham (90°) or active rTMS (0°) group that completed all 20 intervention visits within 5 weeks, with between intervention-visit intervals not exceeding 3 consecutive days.

#### Blinding success

The researcher administering the intervention asked participants to indicate the group they believed they were allocated to, selecting from: real, placebo or unsure. A Likert visual analogue scale (VAS) was used to measure certainty of group allocation after intervention visits 1, 10 or 20. The blinding data presented were collected after intervention visit 1, when it is less likely to be confounded by perceived efficacy,^20, 21^ and successful blinding was defined as >60% ‘unsure’ responses, and approximately even responses across guess categories.

#### MRI

was performed at the Royal Hobart Hospital (GE Optima MR450w 1.5 Tesla scanner). Deidentified baseline and post-intervention scans were analysed and data extracted by researchers at the Sydney Neuroimaging Analysis Centre (SNAC), who were blind to treatment allocation. See supplementary information for MRI acquisition/analysis.

#### PROMs and cognitive / motor assessments

were made at baseline (≤2 weeks preceding or at intervention visit 1), intervention visit 20 (4 weeks [+1] post randomisation) and the 4-months post-randomisation follow-up. PROMs included scores attained on the hospital anxiety and depression scale (HADS), quality of life (AQoL-8D) and fatigue severity scale (FSS). Cognitive and motor assessments included scores attained for the electronic symbol digit modalities test (eSDMT; number of correct matched pair responses), pinching test (number of successful pinches), draw a shape test (tracing error as mean Hausdorff distance across all shapes) and U-turn test (U-turn speed).

### Statistical analysis

Statistical analyses were performed in SPSS (v27, IBM, Armonk, USA), Graphpad Prism (v9). The level for significance was set at p <0.05 (two-tailed). Primary outcome data i.e. treatment-emergent AEs, were not statistically analysed as none were recorded.

The proportion of compliant vs. non-compliant participants per group was compared by a Fisher’s exact test. We did not statistically analyse blinding data, but present summary descriptive statistics, frequency (n) and percentages per guess category, and median certainty scores (VAS), cross-tabulated by intervention group, at each time point.

Continuous MRI outcomes were analysed using MANOVAs and follow-up factorial-mixed ANOVAs. Analyses were run separately for lesion volumes; substructure volumes; myelin-content metrics in peripheral grey matter (MTR and qT1 mapping) or white matter (MTR, qT1 mapping, FA, and MD). ANOVAs included timepoint as the repeated-measures factor and intervention group as the between-subjects factor [sham (90°), rTMS (0°)]. Lobe was included as a repeated-measure, with all cerebral lobes (frontal, parietal, temporal, occipital) included in the lesion volume analyses, but only stimulated lobes (frontal, parietal) included in the peripheral grey-matter and white-matter myelin content analyses. For the white matter myelin content analysis, substructure type (NAWM, lesion) was included as an additional within-subject factor. Significant interactions were identified by simple effects tests that compared timepoints within group and controlled for the false discovery rate (FDR) using the Benjamini-Hochberg procedure (threshold set at 5%). To determine whether participant whole brain volume changed between the baseline and post-intervention MRI, we performed one-sample Wilcoxon signed-rank tests, analysing the SIENA-derived percent whole brain volume change from baseline, to determine whether the percent change was significantly different from 0 (i.e., no change over time) for either group.

Continuous outcomes, including FSS, HADs anxiety and depression, AQoL-8D, U-turn test and draw a shape test, were compared between sham (90°) and rTMS (0°) groups by univariate mixed-ANOVAs. Significant time by intervention interactions were followed-up with simple effects tests, comparing sham (90°) to rTMS (0°) group means within each time point. Outcomes that produced count data, including the e-SDMT and tomato pinching test, were analysed using Mann-Whitney U tests to compare medians in the sham (90°) and rTMS (0°) groups within each time point.

## Results

### Adverse events

Five AEs occurred across the two groups (**Figure 2A**; summarised in **Table 2**), each in a unique participant. Headache and issues related to memory and balance (**Table 2**) were expected AEs.^17^ However, the headache did not occur acutely after the intervention, as would be expected from rTMS^6^ and the ‘issues related to memory and balance’ were reported at the 4-month follow up, by a participant with known variation in these MS symptoms prior to the trial. Therefore, all AEs were classified as unrelated to the intervention by the principal investigator. The single SAE was a participant fall that produced radius and ulna fractures that required hospitalisation and surgery. Following the SAE, the participant was withdrawn from the study, having completed 5 intervention visits. An independent medical monitor evaluated the SAE and deemed it unrelated to the intervention. No participant experienced expected SAEs. No AEs were graded as life-threatening (grade 4) or causing death (grade 5). Except for the AE of ‘issues related to memory and balance’, all AEs and SEAs had resolved by the end of the study.

**Figure 2.**
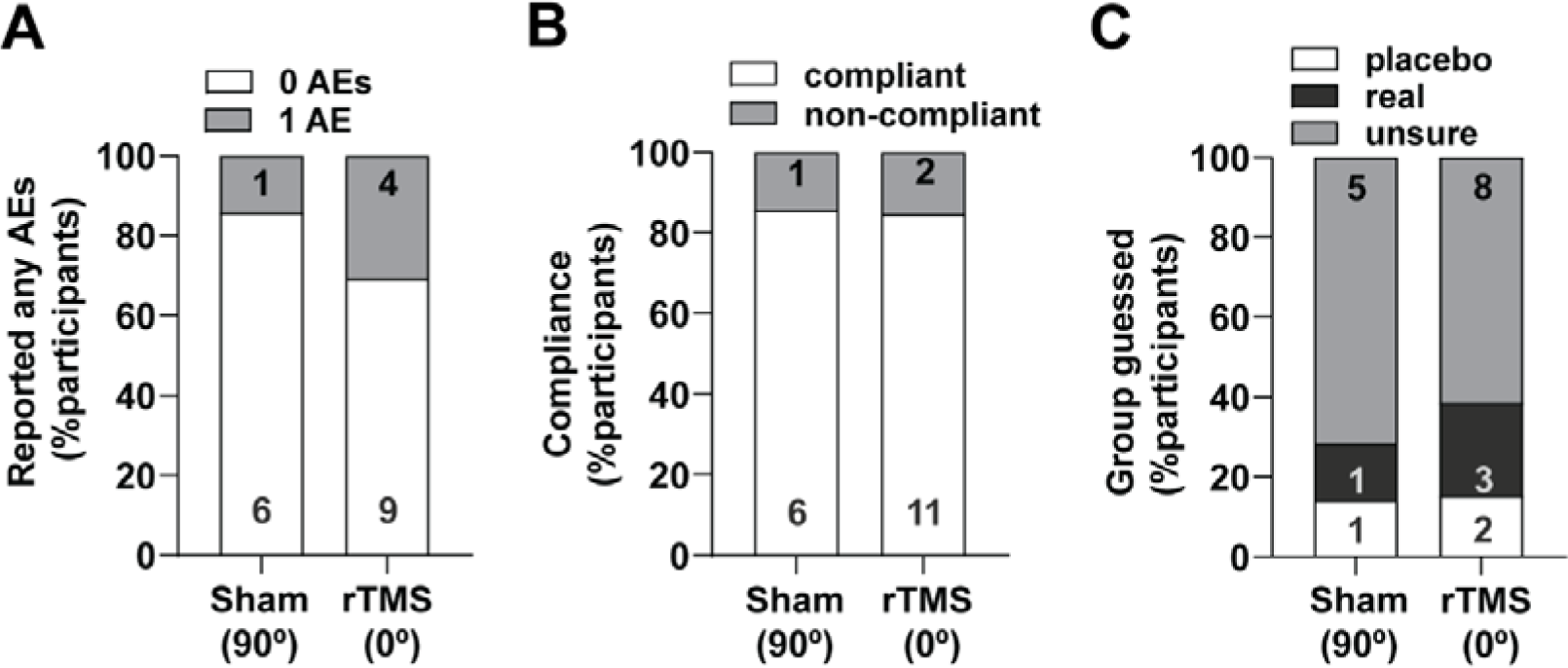
rTMS is safe and tolerable for people with MS. **A)** Proportion (%) and number of participants (within bars) in sham (90°) or rTMS (0°) group that reported no adverse events (AEs, white bar portions), or at least one adverse event (grey bar portions). **B)** Proportion (%) and number of participants (within bars) in sham (90°) or rTMS (0°) groups that were categorised as protocol compliant (white bar portions; defined as completing 20 intervention visits over ≤ 5 weeks with no gaps exceeding 3 days between intervention visits) or non-compliant (grey bar portion). **C)** Proportion (%) and number of participants (within bars) in sham (90°) or rTMS (0°) groups who guessed they were in the placebo (sham) or real group or were unsure, immediately after the first intervention visit.

**Table 2.** Adverse event frequencies and severity by system organ class.

|  | System organ class | MedDRA term(s) | rTMS coil orientation |  | Severity (Grade) | Causality | Expectedness |
| --- | --- | --- | --- | --- | --- | --- | --- |
|  |  |  | Sham (90°)<br>n = 7 | rTMS (0°)<br>n = 13 |  |  |  |
| Any AE n (%) |  |  | 1 (14.3) | 4 (30.7) |  |  |  |
|  | Nervous system disorders | Headache | 0 | 1 (7.6) | Moderate (2) | unrelated | expected |
|  |  | Memory impairment, Ataxia<sup>a,b</sup> | 0 | 1 (7.6) | Mild (1) | unrelated | expected |
|  | Infections and infestations | Shingles | 0 | 1 (7.6) | Moderate (2) | unrelated | unexpected |
|  | Skin and subcutaneous tissue disorders | Other<sup>a,c</sup> | 1 (7.6) | 0 | Mild (1) | unrelated | unexpected |
| Serious AE<sup>d</sup> | Injury, poisoning and procedural complications | Fall, Fracture | 0 | 1 (7.6) | Severe (3) | unrelated | unexpected |

AEs were classified by system organ class and severity, using the common Medical Dictionary for Regulatory Activities (MedDRA), terminology for AEs.<sup>31</sup>
<sup>a</sup> Participant self-reported; clinical assessment unavailable.
<sup>b</sup> Reported at 4-month follow-up, with onset date recalled by participant.
<sup>c</sup> Rash on trunk and hands, possible reaction to Pfizer-Comirnaty COVID-19 vaccination as first dose received within 24 hours of AE onset.
<sup>d</sup> Serious AE requiring hospitalisation. SAEs defined as any AE resulting in death; that is life threatening; requiring hospitalisation or prolongation of hospitalisation; results in persistent disability; consists of congenital abnormality or birth defect; or deemed otherwise medically significant.

### Protocol compliance

85.0% of participants completed 20 intervention visits within 5 weeks, without exceeding the allowed 3 days tolerance interval. For these participants, the intervention period ranged from 25-32 days. One participant in the rTMS (0°) group was withdrawn from the study after visit 5 due to an SAE. Two participants had more than 3 consecutive days between intervention visits, missing 6 or 10 consecutive days due to travel and an unforeseen but mandated quarantine period following travel, respectively. A total of 3 participants were non-compliant, and the proportion of compliant participants was not significantly different between the sham (90°) (6/7) and rTMS (0°) (11/13 compliant) groups (Fisher’s exact test p>0.999; **Figure 2B**).

### Blinding

At the end of intervention visit 1, the proportion (%) of participants who were “unsure” about their group allocation was 71.4% in the sham (90°) and 61.5% in the rTMS (0°) group (**Figure 2C**), consistent with successful blinding. The number and proportion (%) of participants selecting placebo, rTMS or unsure at intervention visit 1, 10 and 20 and the median certainty scores of their guesses are presented in **Table 3**.

**Table 3.**
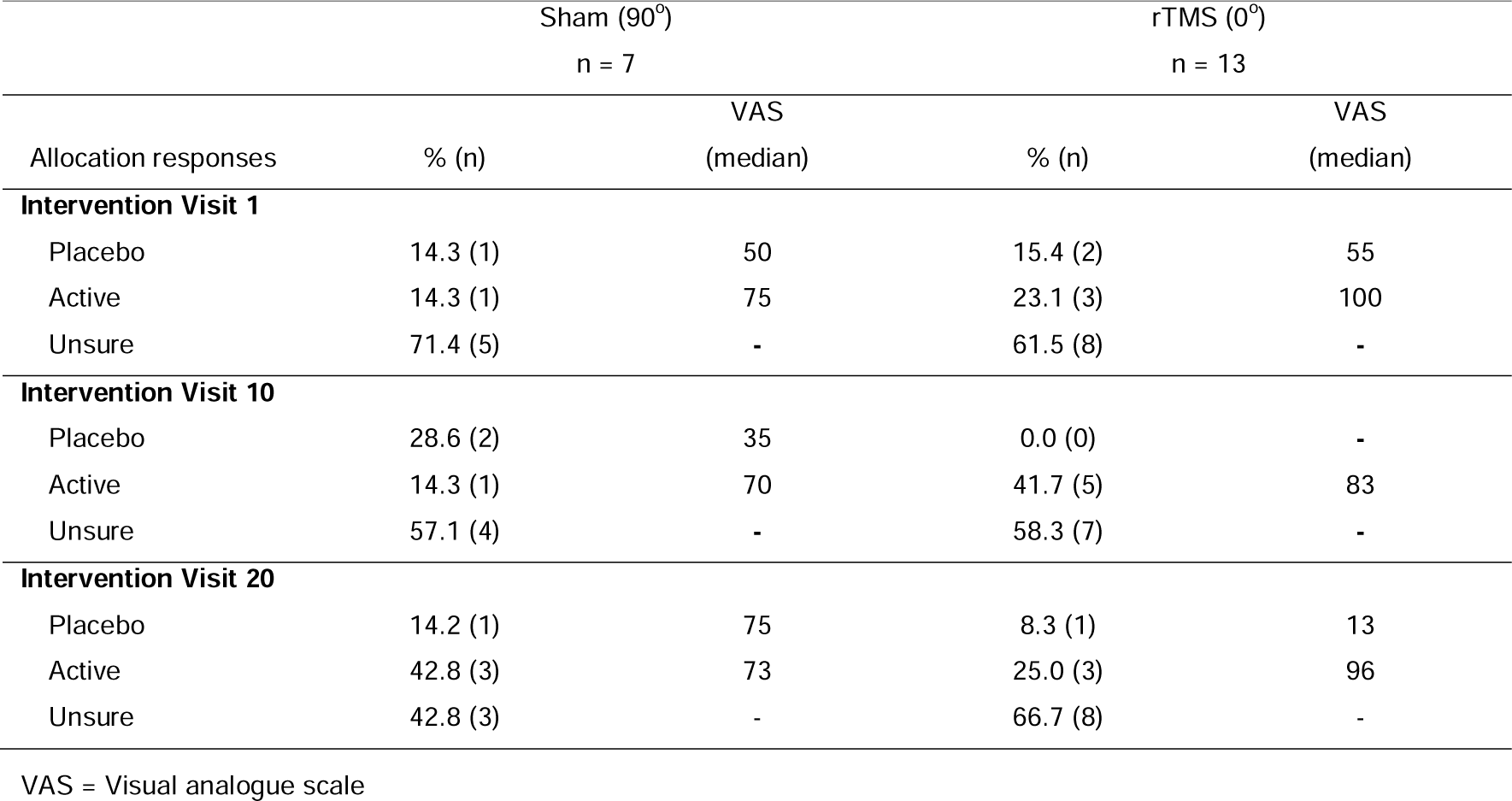
Integrity of blinding at intervention visit 1, 10 and 20 between sham (90°) and active rTMS (0°) groups.

### MRI outcomes

No participant developed new T2 hyperintense lesions (**Figure 3A**) or had lesions enlarge between baseline and follow-up MRIs. Regardless of intervention group, lesion volumes did not change over time across the cerebral lobes (MANOVA, p>0.05), or within frontal and parietal lobes (ANOVAs for each lobe, all p-values >0.05; **Figure 3B**). There was also no evidence of brain atrophy in the sham (90°) or rTMS (0°) groups, as the percentage change in whole brain volume (compared baseline and post-intervention MRIs) was not significantly different from 0 (i.e. no volume change from baseline; one sample Wilcoxon signed-rank tests: sham (90°), p>0.999; rTMS (0°), p=0.233; **Figure 3C**). Furthermore, SIENAX normalised volumes of peripheral grey matter or ventricle cerebrospinal fluid (CSF) did not change (**Figure 3D**; ANOVAs, all p-values >0.05).

**Figure 3.**
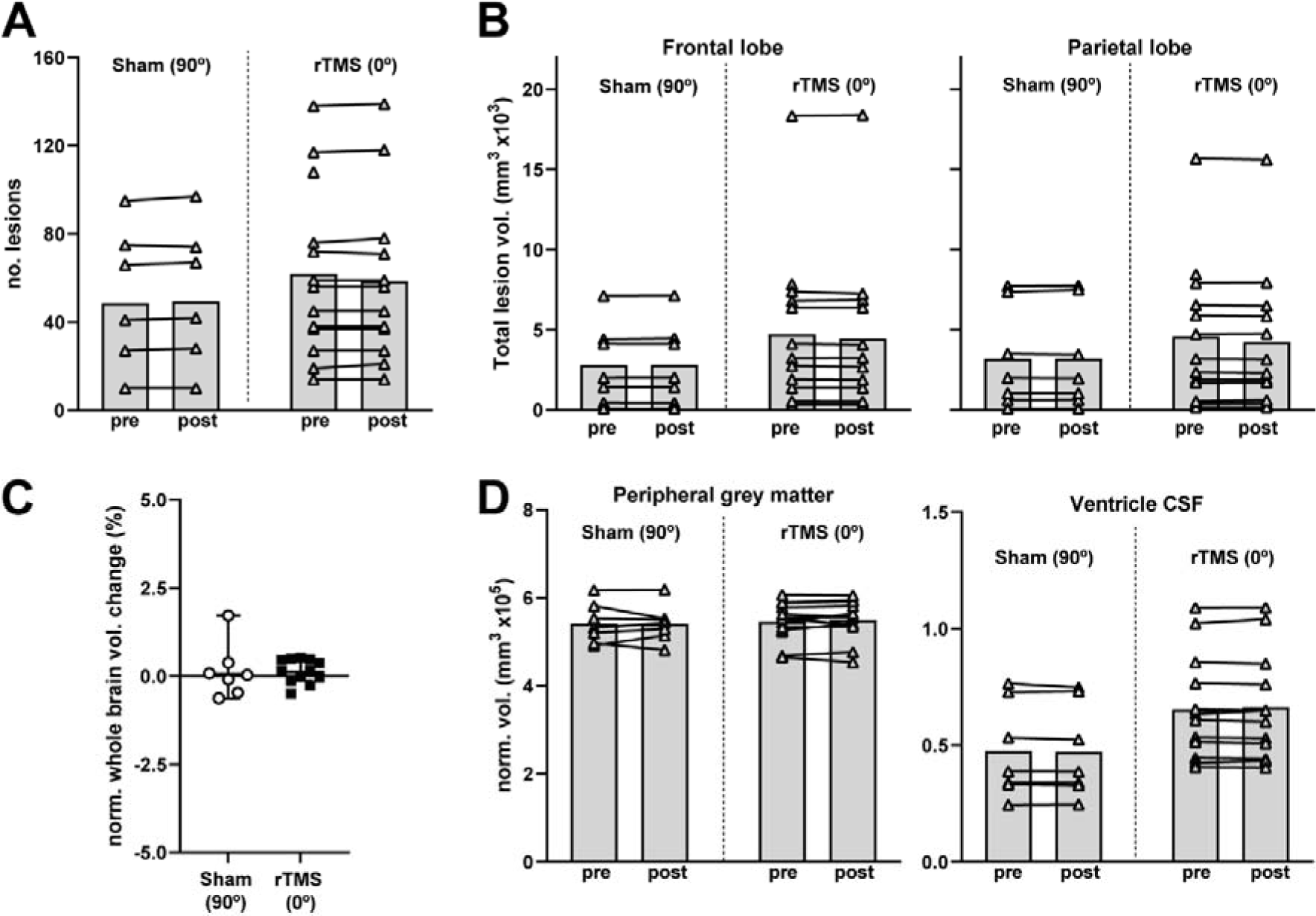
rTMS does not exacerbate MS cortical lesion load or brain atrophy within a 4 to 6-week period. **A)** Number of T2-hyperintense lesions before and after the intervention in the sham (90°) or rTMS (0°) groups. **B)** Lesion volumes in the frontal or parietal lobe before and after the intervention in sham (90°) or rTMS (0°) groups. **C)** Percent brain volume change between baseline and follow-up MRI (lines at the median) in sham (90°) or rTMS (0°) groups. **D)** Normalised volumes of peripheral grey matter or ventricle cerebrospinal fluid (CSF) before and after the intervention in sham (90°) or rTMS (0°) groups.

Across frontal and parietal lobe white matter substructures, MTR, qT1 relaxation rate and FA did not change significantly between timepoints, regardless of group (p-values >0.05, **Figure 4A-F**), but MD changed between timepoints depending on intervention group (time by intervention interaction, p=0.045). MD did not change significantly over time in NAWM, nor in lesions in the sham (90°) group (p-values >0.05). Lesion MD decreased in the rTMS (0°) group but failed to reach significance after FDR correction (p=0.006 > q=0.003, 16 comparisons; **Figure 4G, H**). In peripheral grey-matter, there was a small but significant decrease in mean MTR between timepoints, regardless of intervention group (time p=0.028; all interaction p-values >0.05; **Figure 4I**), while qT1 relaxation rate did not change significantly (all p-values >0.05; **Figure 4J**).

**Figure 4.**
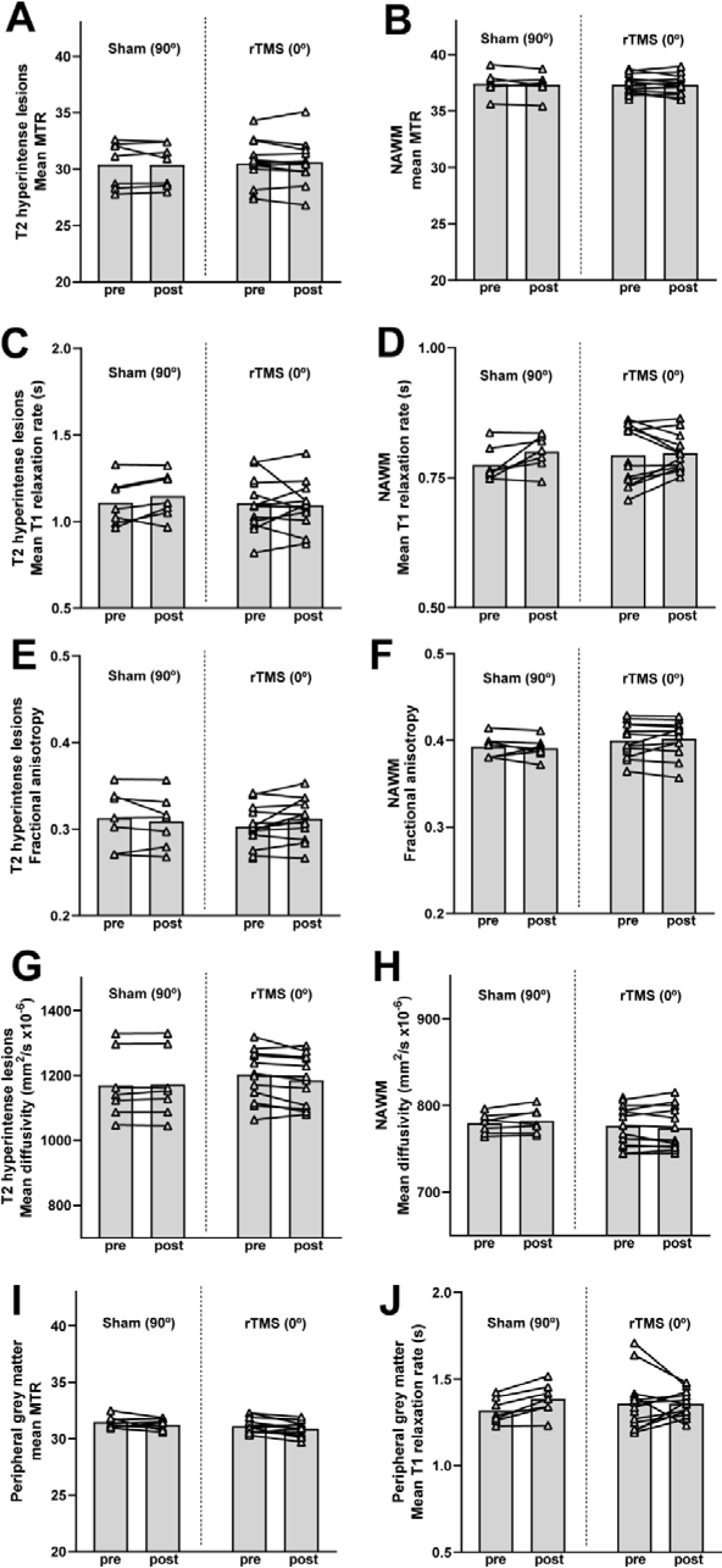
MRI myelin metrics across stimulated frontal and parietal lobes at baseline and after sham (90°) or rTMS (0°) **A)** Mean magnetisation transfer ratio (MTR) of T2 hyperintense lesions. **B)** Mean MTR of normal appearing white matter (NAWM). **C)** Quantitative T1 mapping (qT1) mean relaxation rate of T2 hyperintense lesions. **D)** qT1 mean relaxation rate of NAWM. **E)** Fractional anisotropy in T2 hyperintense lesions. **F)** Fractional anisotropy in NAWM. **G)** Mean diffusivity T2 hyperintense lesions. **H)** Mean diffusivity in NAWM. **I)** Mean MTR of peripheral (cortical) grey matter. **J)** qT1 mean relaxation rate of peripheral (cortical) grey matter. In all graphs, bars represent means; symbols represent means of frontal and parietal lobe values of each participant.

### PROMs and cognitive and motor assessment

Fatigue (**Figure 5A**), anxiety (**Figure 5B**), depression (**Figure 5C**) and overall quality of life (**Figure 5D**) were equivalent for participants in the sham (90°) and rTMS (0°) groups, and did not change significantly between baseline, intervention visit 20 and the 4-month post-randomisation follow-up (all p-values >0.05).

**Figure 5.**
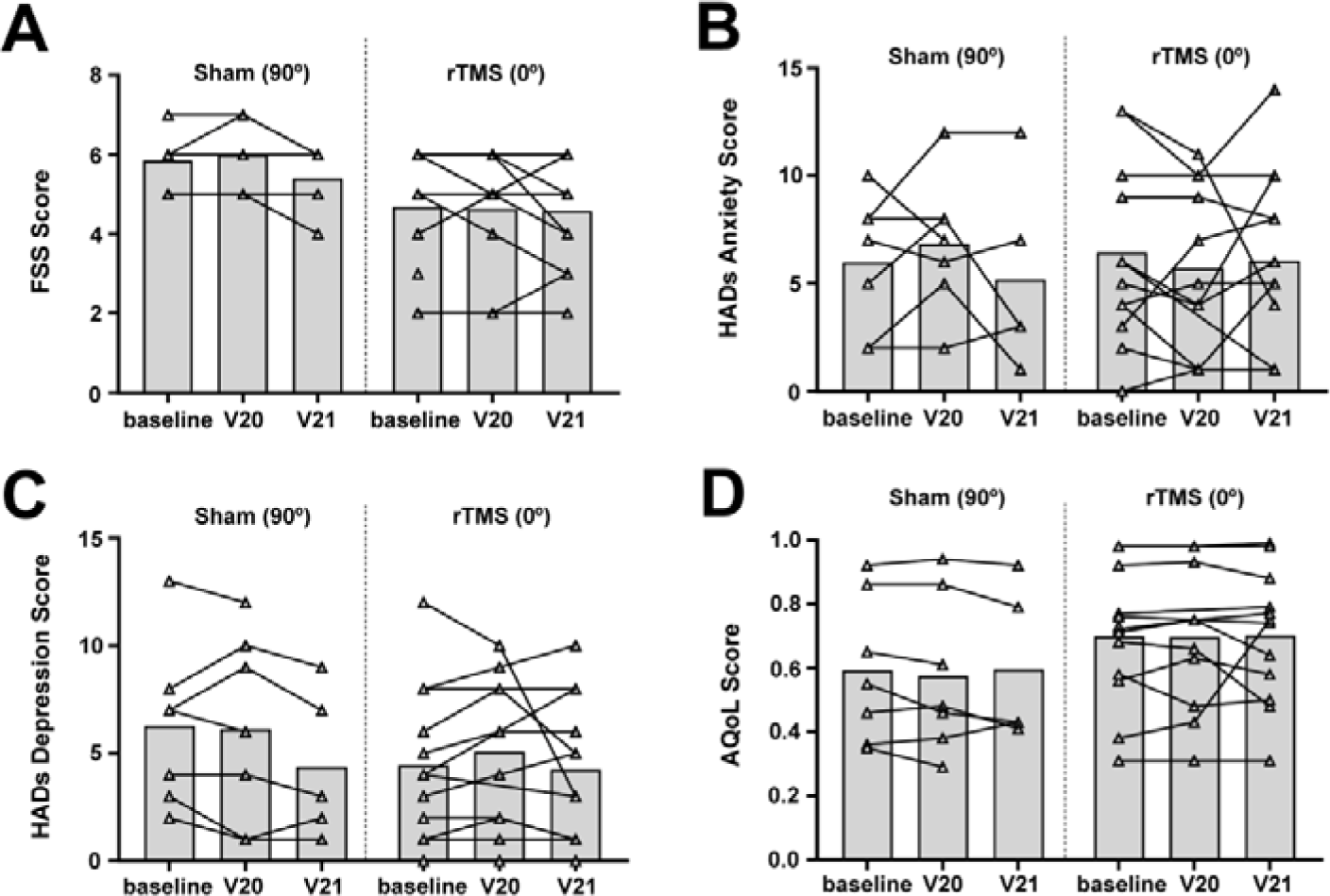
PROMs in the sham (90°) or rTMS (0°) groups at baseline, V20 (post-intervention) and V21 (4-month post-randomisation remote follow-up) **A)** Fatigue Severity Scale (FSS) score. **B)** Hospital Anxiety and Depression Scale (HADs) Anxiety score. **C)** HADs Depression score. **D)** Assessment of Quality of Life 8D score (AQoL-8D).

Data are presented for participants that completed the self-guided cognitive and motor assessments at baseline, intervention visit 20 and the 4-month post-randomisation (**Supplementary table 1**). However, missing, and improbable data points for: the number of correctly matched pairs, the number of successful tomato pinches, tracing errors and average U-turn speed prevented the statistical analyses of these data.

## Discussion

### rTMS is safe for people with MS

Overall, 5 AEs were reported. As the participant retention rate was high (90%-95%) throughout the trial, it is unlikely that AEs went unreported or undetected. None of the AEs were classified as related to rTMS, suggesting that our novel rTMS protocol is safe for people with MS.

### People with MS can feasibly complete 20 rTMS visits

Our rTMS protocol required a substantial time commitment from participants. It is interesting to note that 12/19 (63.2%) participants completed the study according to the ideal rTMS schedule (5 visits per week for 4 weeks). However, 85% of participants were compliant with the protocol, completing all 20 intervention visits within the required 5-week period, with ≤ 3-day interval between intervention visits. At intervention visit 20, participants were invited to provide feedback, and their responses suggest that compliance was enabled by: 1) accommodating appointments around the participants’ schedules anytime between 8am and 6pm and the maximum 3-day interval accommodated long weekends, illness or other factors; 2) free, accessible parking; 3) good communication between participants and researchers, and 4) the gift card provided to offset travel to the study site. These factors, as well as local geography, must be considered when extrapolating from these retention rates to plan a larger study.

As we retained 19 of the 20 randomised study participants at the primary endpoint and 18 out of 20 participants at the secondary endpoint, our study retention rate remained high (90-95%), exceeding the typical cut-off of 80%, which is considered an acceptable retention rate for phase I clinical trials.^22^ No participant voluntarily withdrew, but one was withdrawn from intervention and follow up, at the PIs discretion. One participant was lost to follow up at the 4-month post-randomisation visit and was uncontactable.

### Study blinding and limitations of the sham (90°) intervention

Study blinding was successful as >60% of participants in the sham (0°) and active rTMS (90°) groups reported that they were ‘unsure’ of their group following intervention visit 1. While the sham intervention used here overcomes the bias that would be introduced in an uncontrolled and unblinded trial, we do not recommend it for future efficacy trials. Positioned at 90°, the circular coil delivered rTMS at a lower average intensity than at 0°, and there was a marked difference in the distribution and direction of the induced electric currents, but this was not a true sham comparator. We recommend that future studies include a sham group that positions the coil at 0° (as per rTMS group), reduces the stimulation intensity to 0% maximum stimulator output, and reproduces the sound of rTMS using an auditory recording - allowing the ‘placebo’ effect of trial involvement^23^ to be clearly distinguished from any effect of rTMS on MS clinical outcomes and myelin repair.

### MRI protocols are feasible and provide preliminary evidence of rTMS safety and efficacy

We obtained baseline and post-intervention MRIs for 95% of participants, demonstrating MRI feasibility within our trial protocol. While underpowered, exploratory MRI lesion and atrophy analyses indicate that our rTMS protocol does not exacerbate lesion activity in people with MS, as these within-person measures did not worsen over time in sham (0°) or rTMS (90°) groups. Participants did not develop new lesions and their existing lesions did not enlarge over time. Peripheral grey matter and whole brain volume did not change beyond the expected within-participant variation.^24^

We included multiple MRI metrics that change in response to demyelination and remyelination. When considered together, MD, FA, axial and radial diffusivity are indicative of axonal and myelin integrity.^25^ MTR and quantitative T1 mapping (qT1, mean relaxation time) detect relatively subtle changes in myelin content.^26^ While not significant, the direction of MD change over time is consistent with improved white matter integrity after rTMS. However, any potential change resulting from the intervention was small (∼13um^2^/s), and it is unclear whether white matter changes on this scale reflect a meaningful change in white matter integrity. A larger cohort and the inclusion of: myelin water fractionation, which can detect changes in NAWM myelin over a shorter (3 month) time-course;^27^ SIENAX multi-time-point analyses, to improve detection of longitudinal grey and white matter atrophy, and a 12-month post-randomisation follow-up, would improve an evaluation of the impact of rTMS on MS-relevant MRI metrics.

### It is feasible for participants to complete electronic PROMs; however, the Floodlight app has limitations

There was no change in PROMs outcomes across the time-course or between groups. In an MS population, the FSS has good reliability (ICC = .76, over 6 months) and good-excellent validity metrics.^28^ Neither group altered FSS by ≥ 0.45 points between baseline and 4-month follow-up, which would be considered a clinically-significant change in fatigue.^29^ Importantly, we recorded an 87.5% PROMS data completion rate across timepoints using REDCap marking this as a feasible approach for phase II data collection.

Floodlight cognitive and motor assessments were exploratory in nature, included to assess the feasibility of participants completing these tests within the trial protocol, and were not powered to measure change. Participants experienced technical issues when using the Floodlight Open app in the clinic and at home. They were unable to connect to the internet or experienced data loading or server errors, resulting in 30% of participants failing to complete these measures at the 4-month post-randomisation follow-up; others returned spurious/improbable results. When considering issues of validity^30^ and reliability of data collection, this trial recommends the use of the validated MS functional composite (MSFC) score for functional and cognitive outcome assessments in future trials.

## Conclusions

Our data indicate that 20 sessions of rTMS, delivered in an iTBS pattern over a broad cortical area, is safe, tolerable, and feasible for people with MS. rTMS did not acutely increase lesion load or accelerate brain atrophy over the study period. Following these safety findings, the preliminary efficacy of this rTMS protocol for remyelination will be assessed in people with MS in a randomised, placebo (no stimulation sham) controlled, blinded trial, powered to detect effects on MSFC and MRI metrics of myelin content (TAURUS 2: ACTRN12622000064707).

## Author contributions

PTN: Investigation; methodology; visualization; writing - original draft; writing – review and editing. AZ: Investigation; methodology; visualization; writing – original draft. KM: Formal analysis; data curation; methodology; project administration; supervision; visualization; writing – original draft; writing – review and editing. NS: Software; validation; data curation; methodology; project administration; supervision; writing – review and editing. CE: Formal analysis; data curation; visualization. KK: Investigation; data curation. CW: Investigation; data curation. LL: Investigation; data curation. KDLR: Investigation; project administration. MH: Methodology; funding acquisition; supervision; resources. LJ: Methodology. JR: Conceptualisation; methodology. SC: Investigation. CLC: Conceptualization; funding acquisition; methodology; supervision; writing – review and editing. MB: Methodology; supervision; resources; writing – review and editing. KMY: Conceptualization; funding acquisition; methodology; project administration; resources; supervision; writing – review and editing. BVT: Conceptualization; funding acquisition; methodology; project administration; resources; supervision; investigation; writing – review and editing.

## Data availability statement

Data are available upon request. Please email the corresponding author.

## Data Availability

All data produced in the present study are available upon reasonable request to the authors

## Acknowledgements

We thank Vivienne Jones and the MS Research Flagship’s consumer and community reference committee members for their advice and input. We would like to thank the study coordinators, Chhavi Asthana and Angela McKechnie for support. We also thank Ms. Jennifer Rayner for organisational support of the study at the Clinical Research Facility and Dr. Lauren Giles for providing independent review of SAE. Special thanks to all the study participants.

