## Supplementary files for "Low-intensity repetitive transcranial magnetic stimulation is safe and well tolerated by people living with MS - outcomes of the phase I randomised controlled trial (TAURUS)"

**Supplementary table 1.** Floodlight MS cognition and motor measures in the sham (90^o^) and rTMS (0^o^) at baseline, V20 (post intervention) and V21 (4-month post-randomisation remote follow-up)

|  | Sham (90^o^)  n = 7 | rTMS (0^o^)  n = 13 | Difference between treatment groups | n_1_, n_2_ |
| --- | --- | --- | --- | --- |
| **Five U-Turn Test: Turn speed** | **Mean (95%CI)** | **Mean (95%CI)** | **Mean** |  |
| Baseline visit | 1.264 (1.00 - 1.52) | 1.26 (1.06 - 1.48) | -0.004 (-0.33, 0.32) | 7, 13 |
| Visit 20 | 1.43 (1.16 - 1.69) | 1.34 (1.12 - 1.55) | -0.09 (-0.42, 0.25) | 7, 10 |
| Visit 21 | 1.39 (1.04 - 1.75) | 1.43 (1.19 - 1.68) | 0.04 (-0.39, 0.47) | 4, 7 |
| Change, baseline to Visit 20 | 0.17 (-.010 – 0.42) | 0.07 (-0.15 – 0.29) |  |  |
| Change, baseline to Visit 21 | 0.13 (-0.22, 0.49) | 0.16 (-0.06, 0.41) |  |  |
| **Draw a Shape: Mean Hausdorff Distance Best (lower score more accuracy)** |  |  |  |  |
| Baseline visit | 0.13 (0.09 - 0.16) | 0.13 (0.10 - 0.16) | 0.001 (-0.04, 0.046) | 7, 13 |
| Visit 20 | 0.13 (0.09 - 0.17) | 0.12 (0.09 - 0.15) | -0.01 (-0.05, 0.03) | 7, 11 |
| Visit 21 | 0.06 (0.02 - 0.11) | 0.08 (0.05 - 0.11) | 0.02 (-0.03, 0.07) | 4, 8 |
| Change, baseline to Visit 20 | 0.002 (-0.3 – 0.03) | -0.01 (-0.04 – 0.1) |  |  |
| Change, baseline to Visit 21 | -0.07 (-0.10, -0.02) | -0.05 (-0.07, -0.01) |  |  |
| **Information Processing Speed: Number of correct responses** | **Median (IQR)** | **Median (IQR)** | **Median** |  |
| Baseline visit | 32 (31 - 39) | 35 (27 - 46) | 3 | 7, 13 |
| Visit 20 | 38 (26 - 39) | 43.5 (37 - 50) | 5 | 7, 11 |
| Visit 21 | 33.5 (27 - 41.5) | 45.5 (39 - 51.5) | 12 | 4, 8 |
| Change, baseline to Visit 20 | 5 | 8.5 |  |  |
| Change, baseline to Visit 21 | 1.5 | 10.5 |  |  |
| **Pinching: Number of successful pinches** |  |  |  |  |
| Baseline visit | 25 (22 - 32) | 24 (18 - 34) | -7 | 7, 13 |
| Visit 20 | 33 (17 - 36) | 27 (18 - 34) | -6 | 7, 11 |
| Visit 21 | 30.5 (28 - 33) | 27.5 (16.5 - 33) | -3 | 4, 8 |
| Change, baseline to Visit 20 | 8 | 3 |  |  |
| Change, baseline to Visit 21 | 5.5 | 9.5 |  |  |

Appendix for MRI methodology

| **Hardware** | |
| --- | --- |
| Field strength | 1.5T |
| Manufacturer | GE |
| Model | Optima MR450w |
| Coil type  (e.g. head, surface) | Head |
| Number of coil channels | 8 |

| **Acquisition sequence** | | |
| --- | --- | --- |
| Type  (e.g. FLAIR, DIR, DTI, fMRI) | T2-Weighted FLAIR | |
| Acquisition time | 6.22 minutes | |
| Orientation | Sagittal | |
| Alignment  (e.g. anterior commissure/poster commissure line) | ACPC | |
| Voxel size | 1 × 1 × 1 mm | |
| TR | 5000ms | |
| TE | 116.622ms | |
| TI | 1276ms | |
| Flip angle | 90° | |
| NEX |  | |
| Field of view | 256mm | |
| Matrix size | 256 × 256 | |
| Parallel imaging | **Yes** | No |
| If used, parallel imaging method:  (e.g. SENSE, GRAPPA) | SENSE | |
| Cardiac gating | Yes | **No** |
| If used, cardiac gating method:  (e.g. PPU or ECG) |  | |
| Contrast enhancement | Yes | **No** |
| Type  (e.g. FLAIR, DIR, DTI, fMRI) | T1-Weighted FSPGR |  |
| Acquisition time | 5.43 minutes |  |
| Orientation | Axial |  |
| Alignment  (e.g. anterior commissure/poster commissure line) | ACPC |  |
| Voxel size | 1 × 1 × 1 mm |  |
| TR | 8.48 ms |  |
| TE | 3.248 ms |  |
| TI | 900 ms |  |
| Flip angle | 10° |  |
| NEX |  |  |
| Field of view | 256mm |  |
| Matrix size | 256 × 256 |  |
| Parallel imaging | **Yes** | No |
| If used, parallel imaging method:  (e.g. SENSE, GRAPPA) | GRAPPA |  |
| Cardiac gating | Yes | **No** |
| If used, cardiac gating method:  (e.g. PPU or ECG) |  |  |
| Contrast enhancement | Yes | **No** |
| Type  (e.g. FLAIR, DIR, DTI, fMRI) | Magnetisation Transfer |  |
| Acquisition time | 15.26 minutes (including MT on and off) |  |
| Orientation | Axial |  |
| Alignment  (e.g. anterior commissure/poster commissure line) | ACPC |  |
| Voxel size | 1 × 1 × 3 mm |  |
| TR | 37.0 ms |  |
| TE | 6.0 ms |  |
| TI | - |  |
| Flip angle | 15° |  |
| NEX |  |  |
| Field of view | 256mm |  |
| Matrix size | 256 × 256 |  |
| Parallel imaging | **Yes** | No |
| If used, parallel imaging method:  (e.g. SENSE, GRAPPA) | GRAPPA |  |
| Cardiac gating | Yes | **No** |
| If used, cardiac gating method:  (e.g. PPU or ECG) |  |  |
| Contrast enhancement | Yes | **No** |
| Type  (e.g. FLAIR, DIR, DTI, fMRI) | 16 Direction DTI |  |
| Acquisition time | 9.87 minutes |  |
| Orientation | Axial |  |
| Alignment  (e.g. anterior commissure/poster commissure line) | ACPC |  |
| Voxel size | 2 × 2 × 2 mm |  |
| TR | 16000 ms |  |
| TE | 74.4 ms |  |
| TI | - |  |
| Flip angle | 90° |  |
| NEX |  |  |
| Field of view | 256mm |  |
| Matrix size | 128 × 128 |  |
| Parallel imaging | **Yes** | No |
| If used, parallel imaging method:  (e.g. SENSE, GRAPPA) | ASSET |  |
| Cardiac gating | Yes | **No** |
| If used, cardiac gating method:  (e.g. PPU or ECG) |  |  |
| Contrast enhancement | Yes | **No** |
| Type  (e.g. FLAIR, DIR, DTI, fMRI) | qT1 |  |
| Acquisition time | 8.23 minutes (including both flip angles) |  |
| Orientation | Axial |  |
| Alignment  (e.g. anterior commissure/poster commissure line) | ACPC |  |
| Voxel size | 1 × 1 × 1 mm |  |
| TR | 10.5ms |  |
| TE | 3.7 ms |  |
| TI | - |  |
| Flip angle | 4° & 18° |  |
| NEX |  |  |
| Field of view | 222mm |  |
| Matrix size | 222 × 222 |  |
| Parallel imaging | **Yes** | No |
| If used, parallel imaging method:  (e.g. SENSE, GRAPPA) | GRAPPA |  |
| Cardiac gating | Yes | **No** |
| If used, cardiac gating method:  (e.g. PPU or ECG) |  |  |
| Contrast enhancement | Yes | **No** |

| **Acquisition sequence** | |
| --- | --- |
| If used, provide name of contrast agent, dose and timing of scan post-contrast administration | NA |
| Other parameters: | NA |

| **Image analysis methods and outputs** | |
| --- | --- |
| ***Lesions*** | |
| Type  (e.g. Gd-enhancing, T2-hyperintense, T1-hypointense) | T2-hyperintense |
| Analysis method | Manual Segmentation |
| Analysis software | Jim (Xinapse version 8.0) |
| Output measure  (e.g. count or volume [ml]) | Count and volumes in mm^3^ |
| ***Tissue volumes*** | |
| Type  (e.g. whole brain, grey matter, white matter, spinal cord) | Whole brain, whole brain white matter, whole brain grey matter, cortical grey matter, ventricular CSF |
| Analysis method | Registration based tissue segmentation |
| Analysis software | FSL SIENAX (version 6.0) |
| Output measure  (e.g. absolute tissue volume in ml, tissue volume as a fraction of intracranial volume, percentage change in tissue volumes) | Volumes in mm^3^ normalised for skull size |
| ***Tissue measures (e.g. MTR, DTI, T1-RT, T2-RT, T2*, T2’, ^1^H-MRS, perfusion, Na)*** | |
| **Magnetisation Transfer Ratio (MTR)** | |
| Type  (e.g. whole brain, grey matter, white matter, spinal cord, normal-appearing grey matter or white matter) | Whole brain, whole brain white matter, whole brain normal appearing white matter, cortical grey matter, left/right frontal lobe, left/right parietal lobe, left/right occipital lobe, left/right temporal lobe, left/right frontal lobe white matter, left/right parietal lobe white matter, left/right occipital lobe white matter, left/right temporal lobe white matter, left/right frontal lobe normal appearing white matter, left/right parietal lobe normal appearing white matter, left/right occipital lobe normal appearing white matter, left/right temporal lobe normal appearing white matter, left/right frontal lobe cortical grey matter, left/right parietal lobe cortical grey matter, left/right occipital lobe cortical grey matter, left/right temporal lobe cortical grey matter, whole brain T2-lesion, T2 lesions of left/right frontal lobe, T2 lesions of left/right parietal lobe, T2 lesions of left/right occipital lobe, T2 lesions of left/right temporal lobe |
| Analysis method | Voxel-wise calculation |
| Analysis software | FSL (version 6.0) |
| Output measure | Magnetisation transfer ratio (between 0-100) |
| **Diffusion tensor imaging (DTI)** | |
| Type  (e.g. whole brain, grey matter, white matter, spinal cord, normal-appearing grey matter or white matter) | Whole brain, whole brain white matter, whole brain normal appearing white matter, cortical grey matter, left/right frontal lobe, left/right parietal lobe, left/right occipital lobe, left/right temporal lobe, left/right frontal lobe white matter, left/right parietal lobe white matter, left/right occipital lobe white matter, left/right temporal lobe white matter, left/right frontal lobe normal appearing white matter, left/right parietal lobe normal appearing white matter, left/right occipital lobe normal appearing white matter, left/right temporal lobe normal appearing white matter, left/right frontal lobe cortical grey matter, left/right parietal lobe cortical grey matter, left/right occipital lobe cortical grey matter, left/right temporal lobe cortical grey matter, whole brain T2-lesion, T2 lesions of left/right frontal lobe, T2 lesions of left/right parietal lobe, T2 lesions of left/right occipital lobe, T2 lesions of left/right temporal lobe |
| Analysis method | Voxel-wise calculation |
| Analysis software | FSL (version 6.0) |
| Output measure | FA, MD, AD, RD |
| **Quantitative T1 (qT1)** | |
| Type  (e.g. whole brain, grey matter, white matter, spinal cord, normal-appearing grey matter or white matter) | Whole brain, whole brain white matter, whole brain normal appearing white matter, cortical grey matter, left/right frontal lobe, left/right parietal lobe, left/right occipital lobe, left/right temporal lobe, left/right frontal lobe white matter, left/right parietal lobe white matter, left/right occipital lobe white matter, left/right temporal lobe white matter, left/right frontal lobe normal appearing white matter, left/right parietal lobe normal appearing white matter, left/right occipital lobe normal appearing white matter, left/right temporal lobe normal appearing white matter, left/right frontal lobe cortical grey matter, left/right parietal lobe cortical grey matter, left/right occipital lobe cortical grey matter, left/right temporal lobe cortical grey matter, whole brain T2-lesion, T2 lesions of left/right frontal lobe, T2 lesions of left/right parietal lobe, T2 lesions of left/right occipital lobe, T2 lesions of left/right temporal lobe |
| Analysis method | Voxel-wise relaxometry |
| Analysis software | QUIT (spinicist version 3.3) |
| Output measure | T1 relaxation (ms) |
| ***Other MRI measures (e.g. functional MRI)*** | |
| Type  (e.g. whole brain, grey matter, white matter, spinal cord, normal-appearing grey matter or white matter) | NA |
| Analysis method | NA |
| Analysis software | NA |
| Output measure | NA |

**Other analysis details: NA**
